# Genetic Examination of Hematological Parameters in SARS-CoV-2 Infection and COVID-19

**DOI:** 10.1101/2022.02.28.22271562

**Authors:** Bryce Rowland, Quan Sun, Wanjiang Wang, Tyne Miller-Fleming, Nancy Cox, Misa Graff, Annika Faucon, Megan M. Shuey, Elizabeth E. Blue, Paul Auer, Yun Li, Vijay G. Sankaran, Alexander P. Reiner, Laura M. Raffield

## Abstract

**Background:** People hospitalized with COVID-19 often exhibit hematological alterations, such as lower lymphocyte and platelet counts, which have been reported to associate with disease prognosis. It is unclear whether inter-individual variability in baseline hematological parameters prior to acute infection influences risk of SARS-CoV-2 infection and progression to severe COVID-19.

**Methods:** We assessed the association of blood cell counts and indices with incident SARS-CoV-2 infection and severe COVID-19 in UK Biobank and the Vanderbilt University Medical Center Synthetic Derivative (VUMC SD). Since genetically determined blood cell measures better represent cell abundance across the lifecourse, we used summary statistics from genome-wide association studies to assess the shared genetic architecture of baseline blood cell counts and indices on COVID-19 outcomes.

**Results:** We observed inconsistent associations between measured blood cell indices and both SARS-CoV-2 infection and COVID-19 hospitalization in UK Biobank and VUMC SD. In Mendelian randomization analyses using genetic summary statistics, no putative causal relationships were identified between COVID-19 related outcomes and hematological indices after adjusting for multiple testing. We observed overlapping genetic association signals between hematological parameters and COVID-19 traits. For example, we observed overlap between infection susceptibility-associated variants at *PPP1R15A* and red blood cell parameters, and between disease severity-associated variants at *TYK2* and lymphocyte and platelet phenotypes.

**Conclusions:** We did not find convincing evidence of a relationship between baseline hematological parameters and susceptibility to SARS-CoV-2 infection or COVID-19 severity, though this relationship should be re-examined as larger and better-powered genetic analyses of SARS-CoV-2 infection and severe COVID-19 become available.

## Background

The COVID-19 pandemic caused by the SARS-CoV-2 virus has been responsible for >352 million cases and >5.6 million deaths worldwide as of January 26, 2022 [1]. The clinical course and severity of SARS-CoV-2 infection and illness are heterogeneous. While SARS-CoV-2 infection is characterized by respiratory manifestations and pulmonary complications, infection can elicit a complex immune-inflammatory and thrombotic host response with multi-organ system involvement [2]. Thus, hematologic abnormalities, including T cell lymphopenia, expanded peripheral immature neutrophils, activated monocytes, thrombocytopenia, and lower hemoglobin levels are often observed in hospitalized COVID-19 patients and can fluctuate with disease progression and severity [3-7]. Some of these clinical hematologic laboratory parameters measured at the time of hospital admission have been associated with more severe COVID-19, as well as response to treatment [4, 5]. Further, hematologic, immune, and hemostatic abnormalities may contribute directly to organ damage and dysfunction associated with severe COVID-19 given the central role of blood cells in tissue oxygenation (red cells), innate and adaptive immune response (monocytes and lymphocytes) and thrombosis (platelets, neutrophil extracellular trap or NET formation) [8-14]. Therefore, establishing causal pathways between blood cells and COVID-19 could lead to the discovery of effective treatments for COVID-19 through repurposing existing drugs currently used to treat blood or immune-related disorders [15].

The effect of an individual’s underlying or “baseline” (i.e. before acute infection) hematologic profile on SARS-CoV-2 infection susceptibility and COVID-19 severity is currently not well understood. Previously studies have reported associations between blood cell traits and COVID-19 severity using blood cell indices measured after time of infection or hospitalization, and there have been some inconsistencies across studies [4-7, 16-19]. When blood cell indices are measured after SARS-CoV-2 infection, associations between COVID-19 susceptibility or prognosis with blood cell abundance may reflect acute alterations due to infection or a variety of co-morbidities related to the course of COVID-19 illness (i.e., reverse causality). In order to characterize the relationship between baseline blood cell measurements and risk of SARS-CoV-2 infection or COVID-19 hospitalization, we tested for association between hematological values measured prior to SARS-CoV-2 infection and incident COVID-19 outcomes in two large, longitudinal biobank datasets (UK Biobank and the Vanderbilt University Medical Center Synthetic Derivative (VUMC SD)).

Genetic factors that contribute to hematologic conditions or to inter-individual phenotypic variation in hematologic or immune response parameters may influence host susceptibility or resistance to COVID-19 outcomes [20]. Baseline blood cell measurements are influenced by genetic factors, as well as by long-term environmental, medical, sociodemographic and lifestyle/behavioral factors [21, 22]. Compared to observational studies that assess measured blood cell parameters (either pre-infection or during acute illness), studies of blood cell traits and COVID-19 that leverage genetic variants, which have been constant for a given individual since birth, may be useful in assessing shared genetic architecture and disentangling putative causal relationships. Therefore, we conducted genetic correlation and two-sample Mendelian randomization analyses of blood cell phenotypes with SARS-CoV-2 infection and COVID-19 severity using available summary statistics from the COVID-19 Host Genetics Initiative (HGI) together with a recent GWAS of hematologic traits [23]. Further, we evaluated individual coincident loci from these two analyses. These analyses allowed us to investigate whether genetically determined blood cell parameters measured prior to disease initiation are causally related to COVID-19 susceptibility.

## Results

### Measured Blood Cell Analysis

We tested for associations between baseline-measured levels of 15 hematologic traits and COVID-19 hospitalization and SARS-CoV-2 infection in our discovery data set of 423,358 UK Biobank (UKB) participants (see **Methods, Supplementary Table 1**). Basophil percentage (beta =-0.73, p =3.56e-4) demonstrated association with SARS-CoV-2 infection at the Bonferroni-adjusted threshold (α = 0.05/15=0.003) (**Table 1**). Mean cell hemoglobin (beta = -0.38, p = 2.88e-3), and mean cell volume (β= -0.19, p = 3.49e-4) demonstrated evidence of association with COVID-19 hospitalization at the same Bonferroni-adjusted threshold (**Table 2**). We then attempted to replicate these associations in an independent dataset of up to 1,037,358 participants in the VUMC SD (**Supplementary Table 1**). However, none of these three significant hematology trait-COVID outcome associations observed in UKB were replicated in VUMC SD (**Supplementary Table 2**).

**Table 1.** Measured Blood Cells associated with reported SARS-Cov-2 infection. UKB, UK Biobank; Est., β estimate; s.e., standard error; VUMC SD, Vanderbilt University Medical Center Synthetic Derivative.

| Blood Cell Trait (Units) | UKB Est. (s.e.) | UKB p | VUMC SD Est. (s.e.) | VUMC SD p | Meta-Analysis Est. (s.e.) | Meta-Analysis p | Meta-Analysis Direction |
| --- | --- | --- | --- | --- | --- | --- | --- |
| Basophil (%) | -0.73 (0.2) | 3.56E-04 | 0 (0.17) | 0.996 | -0.31 (0.13) | 2.05E-02 | -- |
| Hematocrit (%) | -0.11 (0.04) | 4.47E-03 | -0.04 (0.02) | 0.01 | -0.05 (0.02) | 7.07E-04 | -- |
| White Blood Cell ($\times 10^9/L$) | 0 (0.08) | 0.96 | -0.12 (0.02) | 4.78E-09 | -0.11 (0.02) | 1.65E-08 | +- |

**Table 2.**
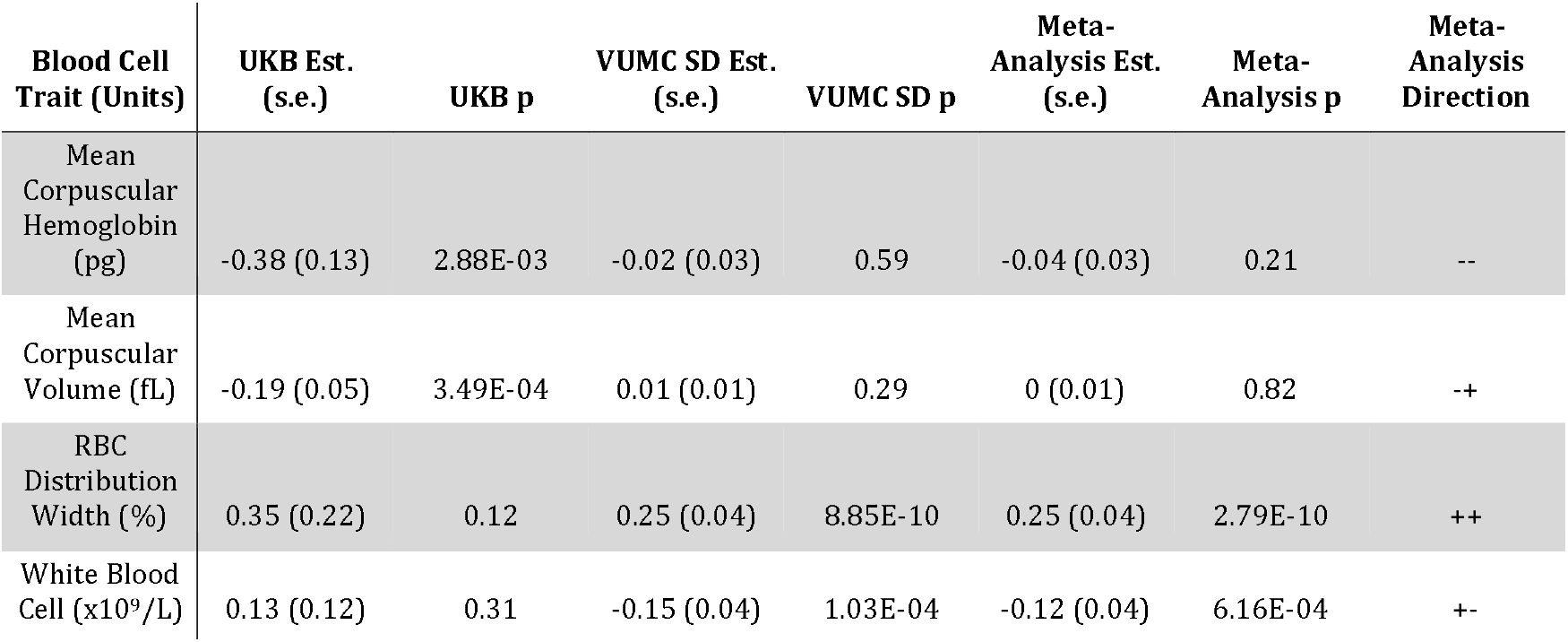
Measured Blood Cells associated with COVID-19 hospitalization. UKB, UK Biobank; Est., β estimate; s.e., standard error; VUMC SD, Vanderbilt University Medical Center Synthetic Derivative.

As a follow-up analysis, we meta-analyzed the measured blood cell – COVID outcome results from UKB and VUMC SD (**Tables 1 and 2**). Total white blood cell count demonstrated evidence of association with both SARS-CoV-2 infection (β = -0.11, p = 1.65 e-8) and COVID-19 hospitalization (β = -0.12, p = 6.16 e-4); however, these associations were primarily driven by the VUMC SD cohort. Additionally, red blood cell distribution width was associated with COVID-19 hospitalization (β = 0.25, p = 2.79 e-10) and hematocrit was associated with SARS-CoV-2 infection (β = -0.05, p = 7.07e-4) (**Tables 1 and 2**)., Given the inconsistencies in results between the VUMC SD and UKB analyses, the remainder of our analyses focus on genetic summary statistic-based analyses, which enable the assessment of invariant factors impacting blood cell abundance across the lifecourse (with similar sample size available for all blood cell phenotypes in well-powered published analyses in UKB [23]).

### Analysis of Coincident Loci for SARS-CoV-2/COVID-19 and Blood Cell Traits

Next, for individual variants previously found to be associated with SARS-CoV-2 infection and COVID-19 severity in recent large meta-analyses from HGI [24], we assessed whether these signals were coincident with a statistically distinct blood cell trait associated variant. If two loci are coincident, this suggests blood cell abundance could be a putative mediator of the SARS-CoV-2/COVID-19 association. We note that such locus level analyses are important even when there is no genome-wide genetic correlation, where differing directions of effect in different regions of the genome, for example, could lead to a null genome-wide result [25]. We obtained summary statistics for conditionally independent GWAS significant variants, i.e. distinct variants, for 29 hematological traits from a recent large GWAS in UKB participants of European ancestry [23]. For each sentinel variant for SARS-CoV-2 infection, COVID-19 severe illness, and hospitalization from the HGI full meta-analysis results, we considered the corresponding lead variant from the meta-analyses excluding UKB (column 1 and 2 of **Supplementary Table 3**). We then assessed coincidence between distinct GWAS variants for blood cell traits from a GWAS in UKB and lead/sentinel variants for both SARS-CoV-2 infection and COVID-19 severe illness and hospitalization from the HGI meta-analysis results [24], based on linkage disequilibrium (LD) (see **Methods**).

Overall, five HGI COVID-19 related sentinel variants were in moderate linkage disequilibrium (r^2^ > 0.4) with at least one distinct blood cell trait variant. Of the five COVID-19 sentinel variants, two are located within highly differentiated regions of the genome with complex patterns of polymorphism, pleiotropy, LD and evolutionary selection (ABO [26, 27] and *HLA* [28]), which would make analysis of coincident signals difficult to interpret and outside the scope of this manuscript. Therefore, we report results on the three remaining COVID-19 sentinel variants based on their associations in: (1) rs4801778 (chr19:49,370,609) which was associated with reported infection and overlaps several red cell-associated GWAS loci; (2) rs74956615 (chr19:10,427,721) which was associated with severe illness and hospitalization and overlaps several GWAS signals for lymphocyte and platelet phenotypes; and (3) rs35081325 (chr3:45,889,921), the sentinel variant for severe illness and COVID-19 hospitalization, which overlaps a nearby locus associated with monocyte count and percentage (**Supplementary Table 3**).

#### Relationship of chromosome 19 (49Mb-50Mb) locus to red blood cell phenotypes

rs4801778, (hg19 position: chr19:49,370,609) is both associated with reported infection in the full HGI meta-analysis and a distinct GWAS variant for reticulocyte count and reticulocyte proportion in UKB. rs4801778 remains the corresponding lead variant at this region after excluding UKB. rs4801778 is in strong LD (r^2^ = 0.935) with rs11541192 (hg19 position: 49,377,424), a missense variant of PPP1R15A (as also noted in [29]). rs11541192 was reported as a distinct GWAS variant for high light scatter reticulocyte proportion in UKB (**Figure 1**). Additionally, rs4801778 is in moderate LD (r^2^ = 0.408) with a second missense variant of *PPP1R15A*, rs556052 (hg19 position: 49,377,436), a distinct GWAS variant for RBC distribution width (RDW). rs4801778 is in moderate LD (r^2^=0.427) with another RDW signal, a 3 prime UTR variant, rs594597. Additionally, our results demonstrate that rs4801778 is in high LD (r^2^ =0.998) with a mean corpuscular hemoglobin variant (MCH), rs3830423 and in strong LD (r^2^ = 0.864) with an immature reticulocyte fraction (IRF) variant, rs73061632 (hg19 position: 49,361,663). Additionally, the frequency of rs4801778 is variable across populations (allele frequency of 16% in African, 19% in European, 14% in South Asian, and 2% in East Asian ancestry populations in 1000 Genomes Phase 3 v5 (1000G)). Our results suggest that the association between SARS-CoV-2 infection and rs4801778 co-localizes with association signals for several red blood cell phenotypes.

**Figure 1.**
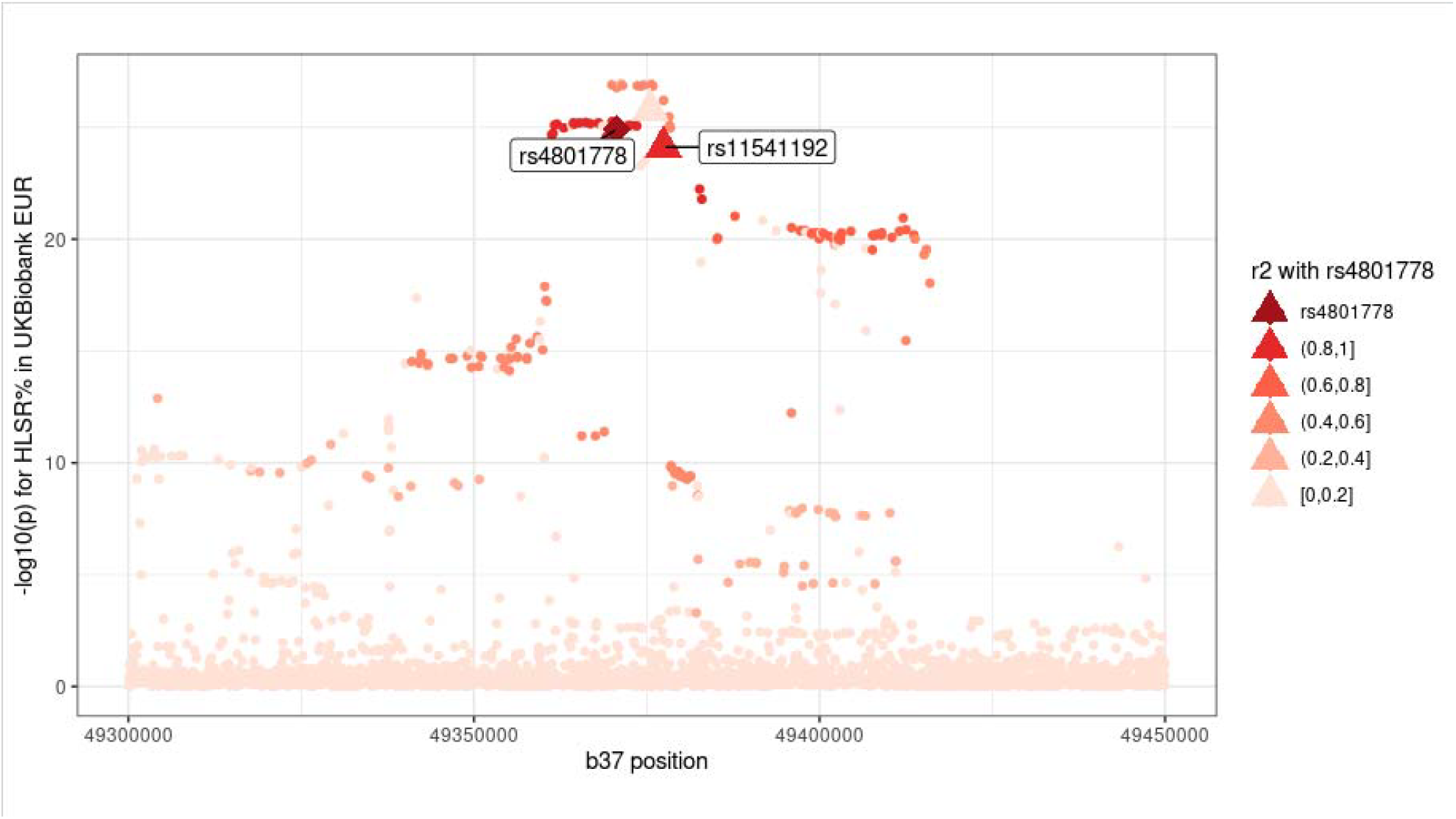
Coincident loci analysis results for rs4801778 and high light scatter reticulocyte proportion in UKB GWAS. rs4801778 (diamond), a lead variant in the HGI COVID-19 GWAS for SARS-CoV-2 infection, was found to be a coincident signal with rs11541192, a distinct variant for high light scatter reticulocyte proportion (HLSR%). rs11541192 is a missense variant for *PPP1R15A*. Triangles are conditionally independent GWAS variants for blood cell traits as determined by conditional analysis in Vuckovic et al. 2020 [23]. Legend: r2 = r^2^.

#### Relationship of chromosome 19 (10Mb-11Mb) locus to lymphocyte and platelet phenotypes

rs74956615 (chr19:10,427,721) was a sentinel variant in the HGI meta-analysis associated with increased risk for severe illness and hospitalization due to COVID-19. After excluding UKB, rs74956615 remained the sentinel variant for severe illness but rs11085727 (chr19:10,466,123) was the sentinel variant for hospitalization. As previously reported [24], rs74956615 is in high LD (r^2^ = 0.75) with rs34536443 (chr19:10,463,118), a missense variant in TYK2. However, rs11085727 is not in LD with rs34536443. rs34536443 was reported as a distinct GWAS variant for lymphocyte count, lymphocyte proportion, platelet count and plateletcrit. Our results suggest that the association between rs74956615 and severe illness and hospitalization due to COVID-19 coincides with lymphocyte and platelet phenotypic associations near *TYK2* (**Figure 2**), suggesting links with immunity and thrombosis related cell types.

**Figure 2.**
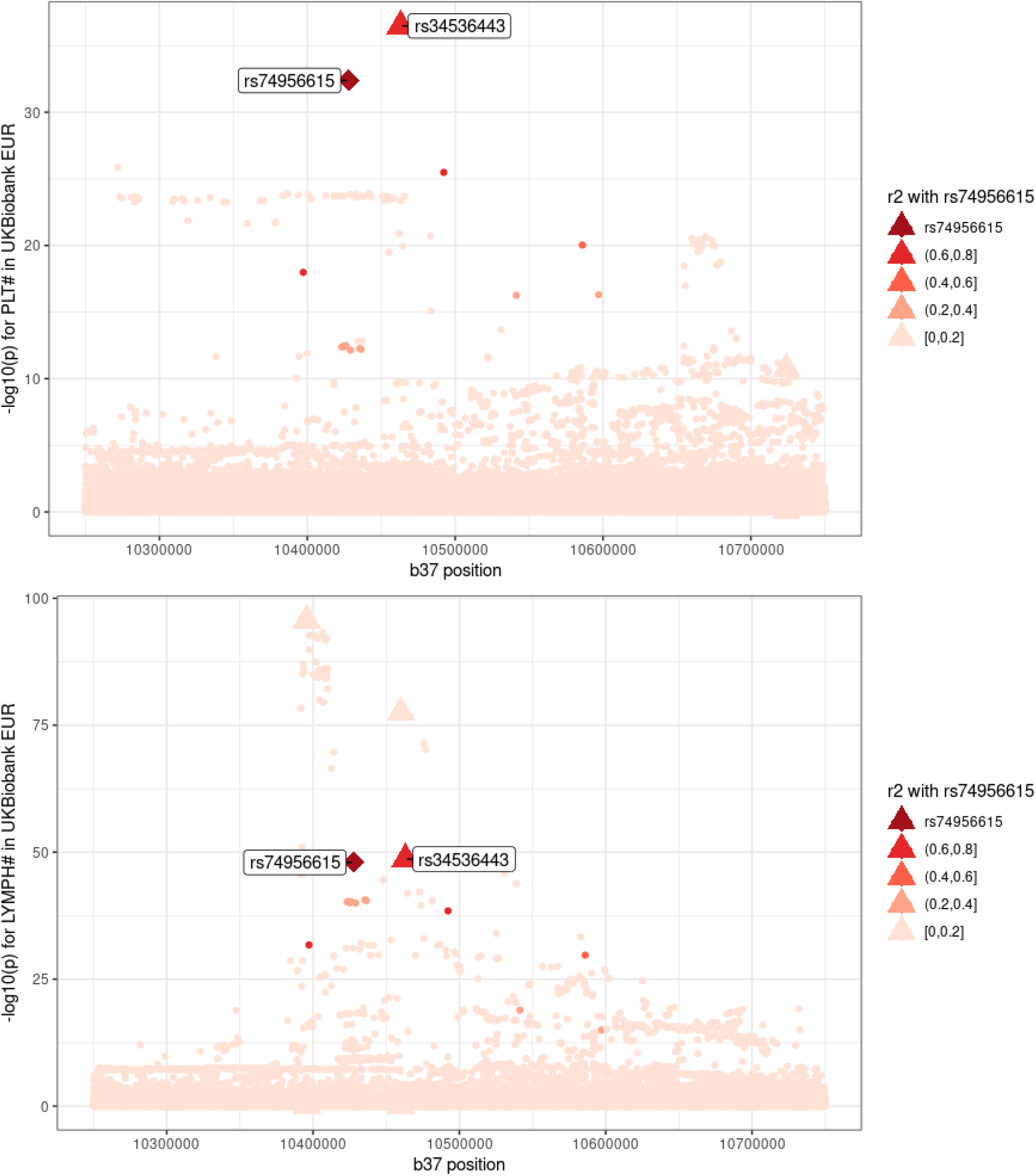
Coincident loci analysis results for rs74956615 and lymphocyte and platelet count in UKB GWAS. rs74956615 (diamond), a lead variant in the HGI COVID-19 GWAS for COVID-19 severe illness and hospitalization, was found to be a coincident signal with rs34536443, a distinct variant for platelet and lymphocyte traits. rs34536443 is a missense variant for *TYK2*. Triangles are conditionally independent GWAS variants for blood cell traits as determined by conditional analysis in Vuckovic et *al*. 2020. Legend: r2 = r^2^.

#### Relationship of chromosome 3 (45Mb-46Mb) locus to monocyte and eosinophil phenotypes

We investigated coincidence between rs10490770 (chr3:45,864,732), a sentinel variant for infection, severe illness, and COVID-19 hospitalization, and nearby monocyte count associations (**Figure 3**). After excluding UKB from the HGI meta-analysis, rs35081325 (chr3:45,889,921) is the lead variant for severe illness and hospitalization, and rs35508621 (chr3: 45,880,481) is the lead variant for SARS-CoV-2 infection.

**Figure 3.**
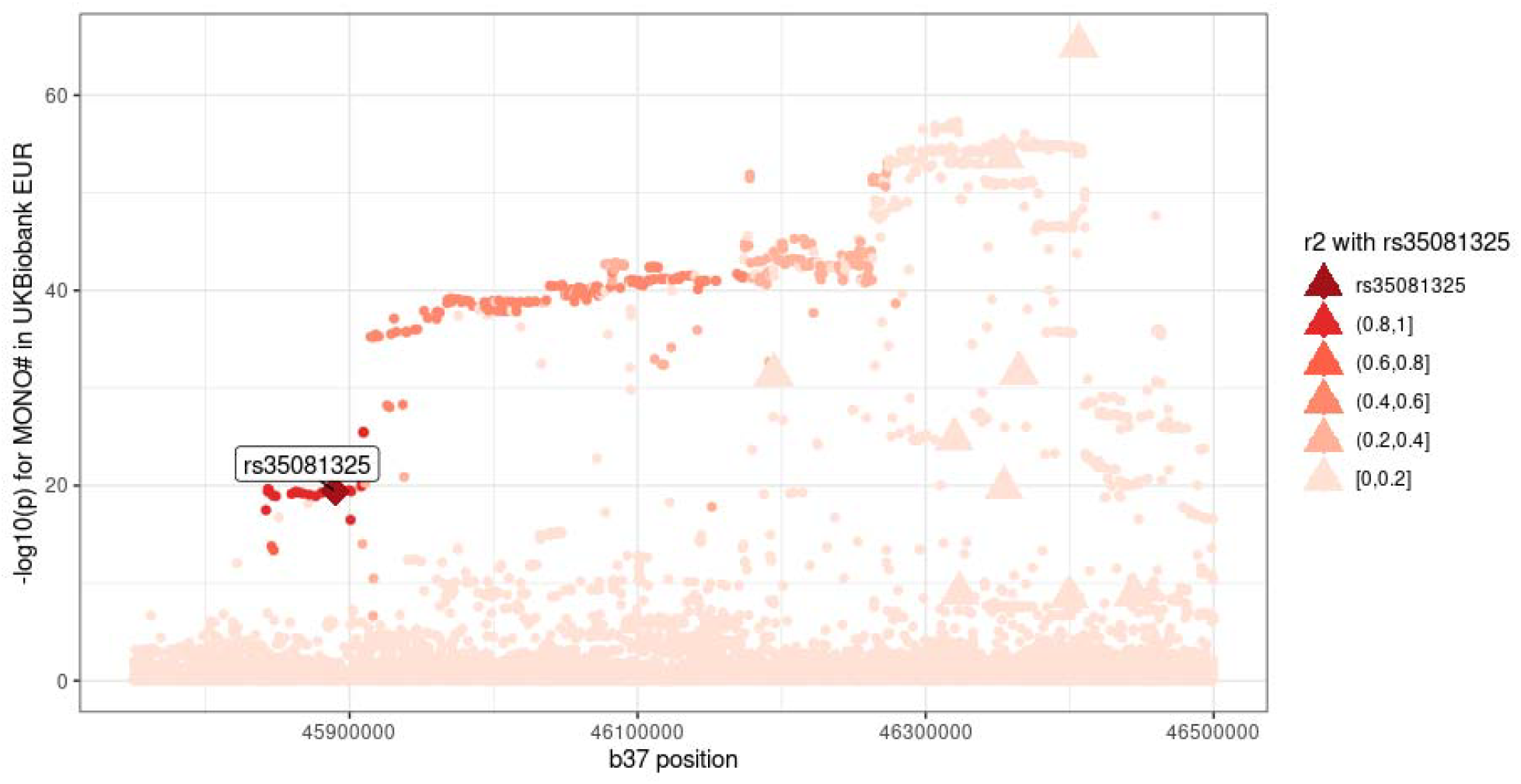
Coincident loci analysis results for rs35081325 and monocyte count in UKB GWAS. rs35081325 (diamond), a lead variant in the HGI COVID-19 GWAS for COVID-19 severe illness and hospitalization was not found to be coincident with any of the nine nearby monocyte GWAS distinct variants. Triangles are conditionally independent GWAS variants as determined by conditional analysis in Vuckovic et al. 2020. Legend: r2= r^2^.

A previous GWAS of hematologic traits identified nine distinct monocyte count GWAS variants within 2Mb of the COVID sentinel variant(s). Of the nine distinct monocyte GWAS variants, none were in LD with rs10490770 or rs35508621 (LD r^2^ <= 0.05). However, both rs35081325 and rs35508621 are in moderate LD (r^2^ = 0.47 with both variants, respectively) with rs74586549 (chr3:45,926,043), a *LZTFL1* intronic variant which was identified as a distinct GWAS signal for eosinophil percentage (**Figure 4**) [23]. Additionally, the frequency of both variants are highly variable across populations (minor allele frequency of 0.4% in African, 8% in European, 30% in South Asian and 0.5% in East Asian populations in 1000G). Our results do not provide evidence of coincidence with monocyte associations but do suggest eosinophils as a potential mediator of this COVID-19 severe illness and hospitalization associated locus. Analyses in this region may benefit from future fine mapping studies with larger sample sizes in diverse ancestral populations.

**Figure 4.**
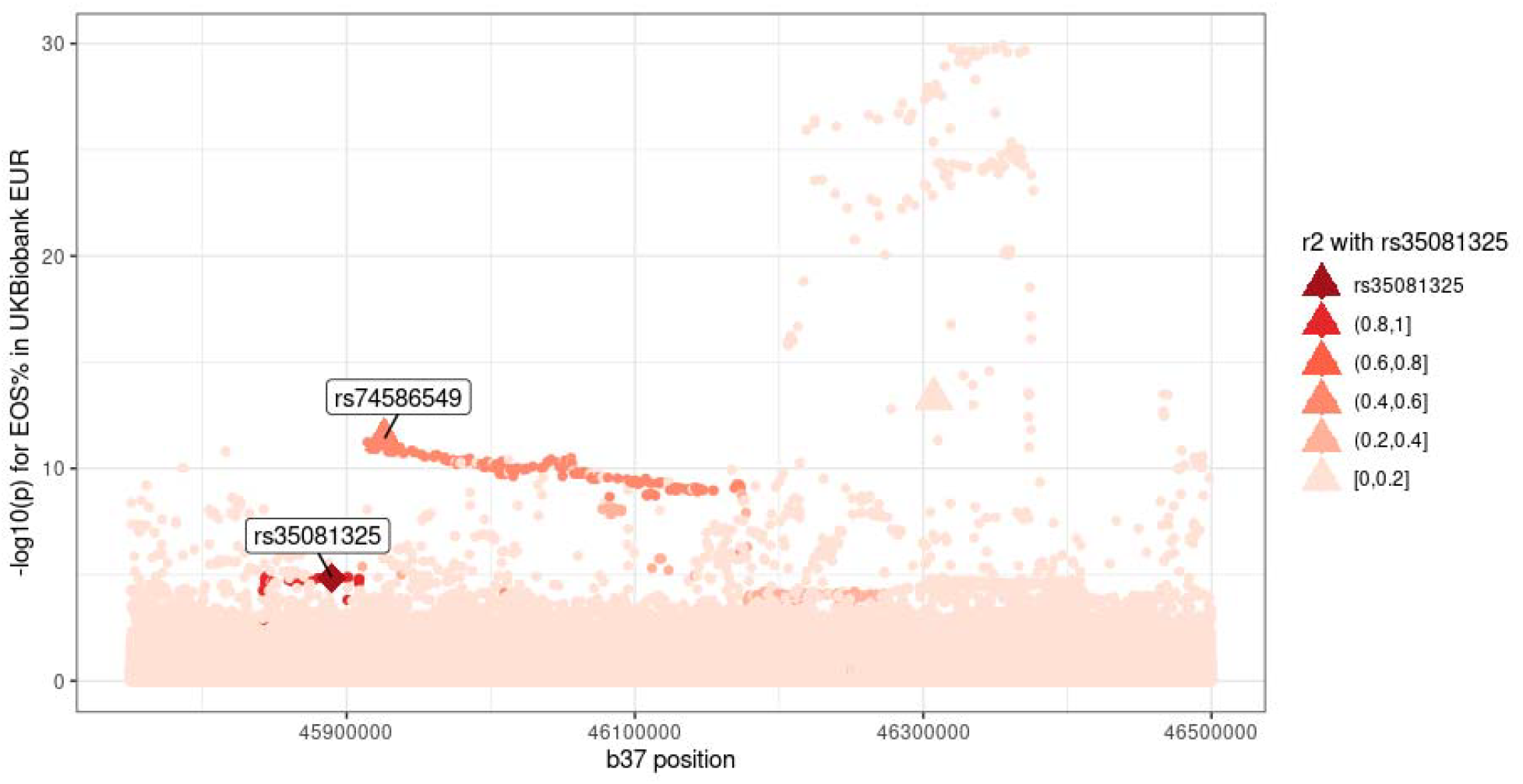
Coincident loci analysis results for rs35081325 and eosinophil percentage in UKB GWAS. rs35081325 (diamond), a lead variant in the HGI COVID-19 GWAS for COVID-19 severe illness and hospitalization was found to be coincident with rs74586549, a distinct GWAS variant for eosinophil proportion. Legend: r2 = r^2^.

### Genetic Correlation Analyses

In order to assess the shared genetic architecture of blood cell traits with COVID-19 severity and SARS-CoV-2 reported infection, we performed LD Score Regression to estimate genetic correlation. Genetic correlation provides an estimate of the overlap in terms of evidence of associations for a pair of complex traits with existing genome-wide summary statistics [30]. Overall, we estimated genetic correlations for 29 blood cell trait phenotypes with reported SARS-CoV-2 infection, severe COVID-19 illness, and hospitalization.

There were no significant genetic correlations at the Bonferroni-corrected threshold (α = 0.05/87) (**Supplementary Table 4**). We did identify two nominally significant (p < 0.05) genetic correlations: mean sphered cell volume with hospitalization, and immature reticulocyte fraction with COVID-19 severe illness (**Table 3**). There were no nominally significant genetic correlations between blood cell traits and reported infection (**Supplementary Table 4**). Of all the blood cell traits tested, lymphocyte percent had the smallest p-value with reported infection (*rg* = 0.138, p = 0.1051).

**Table 3.** Significant LD score regression genetic correlation estimates, standard errors and p-values.

| COVID-19 phenotypes | Blood Cell Trait | Genetic Correlation Estimate | Genetic Correlation se | p-value |
| --- | --- | --- | --- | --- |
| Hospitalization | Mean Sphered Cell Volume | 0.09 | 0.04 | 0.04 |
| Critical illness | Immature Reticulocyte Fraction | 0.10 | 0.05 | 0.05 |

### Mendelian Randomization Analyses

In order to further assess shared genetic architecture as well as potential directional causal associations between blood cell traits measured pre-infection and the three COVID-19 outcomes, we performed Mendelian randomization (MR) analysis with blood cell traits as exposures and COVID-19 phenotypes as outcomes. In contrast to genetic correlation and analysis of coincident loci, MR analyses attempt to assess the causal effect of one trait on another, not simple local or genome-wide sharing of genetic association signal [25]. Our instrumental variables were the distinct variants identified from previous GWAS signals for blood cell traits in individuals of European ancestry, as used above for analysis of coincident loci (see **Methods**; [23]). Compared to the instrumental variables used in the MR analysis in the HGI flagship paper (all GWAS sentinels for an exposure trait), our reduced set of distinct variants is less likely to contain weak instruments that would bias the MR causal estimate [31, 32]. Additionally, samples were excluded from the hematological trait GWAS in UKB European ancestry participants based on relevant exclusions to blood cell variation (such as excluding individuals with positive pregnancy status, certain drug treatments, etc, **Supplementary Table 6**), rather than including all UKB European ancestry participants. For our analysis, we used the HGI COVID-19 summary statistics excluding UKB participants[33] to prevent potential confounding. When there is no significant evidence of directional pleiotropy (MR-Egger Intercept p-value > 0.05) we report the causal effect estimate as the inverse-variance weighted (IVW) estimator of the causal effect on the log odds ratio scale. Otherwise, we report the MR-Egger causal effect estimate. As a measure of robustness, we provide the weighted median estimates of the causal effects as well. The full MR results are shown in **Supplementary Table 5**.

There was only one significant MR test at a stringent Bonferroni-adjusted threshold (α = 0.05/87). We found that variants affecting basophil proportion also affect COVID-19 hospitalization (MR-Egger causal estimate 7.84, p = 7.8e-8), with significant directional pleiotropy effects (MR-Egger Intercept p = 6e-7). In addition, we identified one hematologic trait with a nominally significant MR test (p < 0.05) with COVID-19 hospitalization, and one trait with reported infection (**Table 4**). Specifically, variants associated with increased mean platelet volume are associated with increased risk of COVID-19 hospitalization (OR = 2.067, p = 8.7e-3) (**Figure 5**), and variants affecting red blood cell count have a negative effect on reported infection (OR = 0.412, p = 0.017).

**Table 4.** Significant Mendelian randomization causal estimates.

| COVID-19 Traits | Blood Cell Trait | IVW causal estimate | IVW p-value | MR Egger intercept estimate | MR Egger intercept p-value | MR Egger causal estimate | MR Egger causal p-value |
| --- | --- | --- | --- | --- | --- | --- | --- |
| Hospitalization | Basophil (%) | 0.99 | 0.37 | -0.19 | 5.98E-07 | 7.84 | 7.84E-08 |
|  | Mean Platelet Volume | 0.73 | 8.70E-03 | 0.03 | 0.14 | 0.20 | 0.65 |
| Reported infection | Red Blood Cell Count | -0.16 | 0.41 | 0.03 | 0.02 | -0.89 | 0.01 |

**Figure 5.**
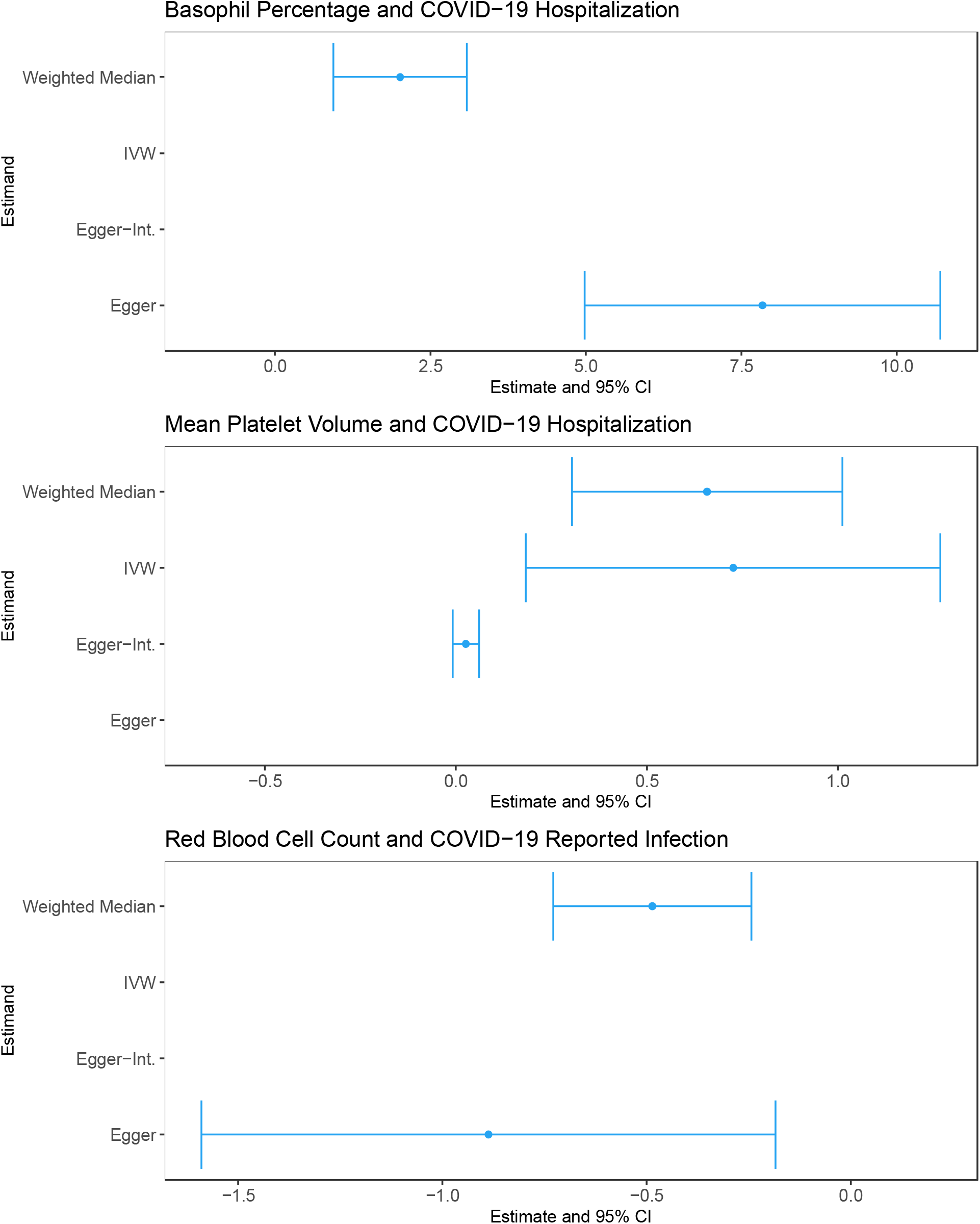
Selected Mendelian randomization results. Each subfigure shows the estimates (denoted by dots) and their 95% confidence intervals (denoted by the range of each bar) for four different estimands. Blue color represents significant associations.

## Discussion

Given the extensive and variable reports that altered blood cell phenotypes are found in the setting of COVID-19 illness following SARS-CoV-2 infection, we studied both associations between baseline blood cell traits and COVID-19 outcomes as well as potential causal relationships with blood cell traits as exposures for COVID-19 outcomes via MR. Our results do not support a clear role of baseline blood cell traits (prior to SARS-CoV-2 infection) in risk of SARS-Cov-2 infection or COVID-19 hospitalization, though we argue our genetic analyses in particular should be repeated in the future as sample sizes accumulate for COVID-19 related GWAS analyses.

The mostly modest hematologic trait associations with COVID-19 outcomes we observed in the UKB (ex., basophil percentage and reported SARS-CoV-2 infection) could not be replicated in the VUMC SD cohort, and had little support from the existing literature on blood cell indices measured at time of infection/hospital admission. We hypothesize the inconsistencies between the UKB and VUMC SD cohorts may be due to multiple factors. Notably, UKB and VUMC SD used differing methods of selecting individuals for blood cell measurement (with UKB measuring blood cell indices at a single timepoint in all participants with a baseline blood sample, and VUMC SD using median values across multiple blood cell measurements, and blood cells only measured in a subset of participants, with this subsetting based on a clinical need to run hematological assays). There are also different cohort characteristics across these two biobanks (**Supplementary Table 1**), with differing sample sizes for different blood cell traits also potentially contributing in VUMC SD (**Supplementary Table 2**). The somewhat more compelling associations in VUMC SD alone of lower white blood cell counts with higher odds of reported SARS-CoV-2 infection (perhaps reflecting immune suppression) and of higher RDW with higher odds of COVID-19 hospitalization (concordant with previous associations of RDW at time of COVID-19 hospitalization with mortality [34], as well as prior RDW associations with clonal hematopoiesis [35], inflammation[36], and mortality in other contexts [37, 38]) were not replicated in UKB. Lack of replication may in part be due to systematic differences in biobank recruitment (notably clinical ascertainment for VUMC SD, with nonrandom reasons for which blood cell assays are ordered in this clinical setting, versus population-based sampling in UKB across a narrow age range, with a recruitment bias towards healthier participants than the UK population as a whole [39]). By contrast, blood cells might be measured more frequently in VUMC SD in participants impacted by certain diseases or using certain medications. Such differences between populations can lead to variability in measured phenotypes and highlights the value of examining genetic variation underlying such traits, though of course genetic findings can also be susceptible to gene x gene and gene x environment impacts that may vary by population.

Given the extensive data we have on genetic variants impacting blood cell traits [23, 40, 41], we next attempted to examine both coincidence of specific loci associated with blood cell traits and COVID-19 susceptibility/severity and genome-wide summary-based measures, including overall genetic correlation and MR analyses. Our analyses demonstrate a lack of concordance between MR and genetic correlation results and the strongest individual coincident associations with measured blood cells (ex., the *PPP1R15A* locus on chr19p13 and red blood cell parameters). The measured blood cell traits that are most strongly associated with COVID-19 related hospitalization in the combined VUMC SD and UKB analysis, including RDW, may be tagging more general inflammatory pathways as opposed to playing a causal role in disease pathogenesis, based on the nonsignificant MR results. We do note that these analyses should be repeated with future iterations of the SARS-CoV-2/COVID-19 genetic analyses, and nominal overall relationships such as MR for red blood cell counts and infection (supported by coincident individual locus on chr19p13) should be further examined in future releases of COVID-19 focused genetic association summary statistics. We note that the heritability estimates of severe illness and hospitalization for COVID-19 were small (h^2^ = 2.8e-3, se = 7e-4 and h^2^ = 1.9e-3, se = 5e-4, respectively), as was the estimated heritability of reported infection (h^2^ = 9e-4, se = 3e-4), consistent with estimates reported in the HGI flagship manuscript [29]. By contrast, we found that the 29 assessed blood cell traits had estimated heritabilities of up to 0.31 in our genetic correlation analyses (ranging from 0.04 to 0.31). Our analyses are thus likely underpowered due to low heritability explained by existing COVID-19 related GWAS. Analysis of significance trajectories suggests many more COVID-19 related genetic loci remain to be identified, including potentially immune related loci which may be relevant to blood cell traits [42].

Based on analysis of coincident genetic loci, a genomic region on chromosome 19p13 harboring one of the index variants for SARS-CoV-2 infection overlaps an association signal for several RBC traits including reticulocyte count and proportion (and partially overlaps association signals for RDW). The red cell – SARS-CoV-2 infection association signal in this region overlaps two genes, *PLEKHA4* (encoding pleckstrin homology domain containing A4) and *PPP1R15A* (protein phosphatase 1 regulatory subunit 15A). As described under **Results**, this association signals includes several missense variants of *PPP1R15A*. The protein encoded by *PPP1R15A* is also known as GADD34 (Growth arrest and DNA damage-inducible protein 34) and is highly expressed in bone marrow. In mice, knockout of Gadd34-is associated with decreased erythrocyte volume, increased numbers of circulating erythrocytes, and decreased hemoglobin content, resembling human thalassemia syndromes, due to the reduced initiation of the globin translation machinery [43]. PPP1R15A/GADD34 activity is also induced following cellular stress and has been implicated in response to viral infections (including coronavirus) and type 1 interferon production in humans and other species [44-48]. Finally, as noted above, there are strong differential allele frequencies of the sentinel *PPP1R15A* variant across global ancestral populations, a pattern that has been reported for other COVID-19 host susceptibility variants [49, 50]. Infection-related variants have been found for multiple viruses in regions under selective pressure or with allele frequency differences across populations [51]. As sample sizes increase for COVID-19 related genetic analysis, it will also be interesting to examine the associations of red blood cell related variants with COVID-19 severity, given potential links with oxygenation/clotting during disease progression.

As noted previously[29], the COVID-19 associated sentinel variant rs7495661 and its LD surrogate, the *TYK2* missense variant rs34536443 (p.Pro1104Ala), have been previously associated with risk of various autoimmune diseases [52], higher lymphocyte count, and lower platelet count. *TYK2* is involved in immune response in humans and *TYK2* deficiency results in impairment of cytokine response in mouse models [53, 54]. Given the role of *TYK2* in host autoimmunity, it is possible that the association with lower platelet count may represent an autoimmune phenomenon due to autoantibodies that inhibit platelet production.

Recently Sun *et al*. reported an MR analysis of WBC phenotypes [55] using the set of distinct genetic variants from Vuckovic et al. [23] and Chen *et al*. [40] Our variant sets differ slightly from those in Sun *et al*. due to their incorporation of distinct signals from the multi-population analysis of blood cell traits and the analysis of the multi-population HGI summary statistics with larger sample size. Inclusion of multi-population summary statistics may lead to issues with the validity of genetic correlation and MR methods, due to differences in linkage disequilibrium patterns across global populations and across different multi-population summary statistic sets. Nonetheless, those results are consistent with ours with respect to direction and effect size for basophils and WBCs. Similarly, Wang *et al*. also performed a MR analysis for hematological parameters and severe COVID-19, but using an earlier freeze of the HGI summary statistics and without consideration of multiple testing or of the distinct signal list derived in Vuckovic *et al*. by direct conditional analysis (SNP selection was instead based on LD clumping using the 1000G Phase 3 reference panel). [56]

We would highlight several key limitations in this work. First, the time between SARS-CoV-2 infection/COVID-19 hospitalizations and blood cell measurements were variable among individuals and different in UKB. We have attempted to account for this, but also emphasize that external factors (diseases, diet changes, *etc*) may alter hematopoiesis and measured blood cell counts, in contrast to genetic factors associated with these traits, where alleles are assigned at birth. Second, the measures we examined here are those from readily obtained peripheral blood cell counts, but there are other interesting hematological measures which are less frequently assessed in large populations. For example, specific lymphocyte subsets (such as naïve or memory cells) may be relevant to COVID-19 but could not be assessed here. Third, the power of MR analysis is still limited for COVID-19 genetics, even with the coordinating efforts of the HGI meta-analysis. We conducted a MR power analysis at (α = 0.05/87) using the case and control counts from the HGI meta-analysis and the minimum and maximum estimated heritability from the LD Score Regression analysis for our blood cell traits (h^2^ range: 0.04 -0.3, **Supplementary Figure 1**). The limited heritability explained by the variants identified in the HGI meta-analyses causes challenges in using both genetic correlation and MR analysis methods. There is a more up to date version of the HGI meta-analysis which includes more samples (Release 6 [57]), but European-ancestry-only summary statistics and summary statistics excluding UKB are not yet available. Fourth, COVID-19 case/control definitions in available GWAS summary statistics are limited by lack of specificity (ex., the use of population controls and lack of more specific information about disease course), though such specificity trade-offs are common to maximize power for case/control phenotypes. Finally, we note that while several blood cell trait-associated loci coincide with signals for COVID-19 severity and SARS-Cov-2 infection, it is difficult to formally evaluate enrichment, given the high polygenicity and large number of genome-wide significant signals known for highly heritable blood cell phenotypes.

## Conclusion

Despite the strong epidemiological links with blood cell indices and COVID-19 related phenotypes in individuals with existing SARS-CoV-2 infection, as well as the reasonable putative biological relevance through roles in immunity, oxygenation, and thrombosis, we see little conclusive evidence of association between pre-infection blood cell indices and incident SARS-CoV-2 infection and COVID-19 severity, using either epidemiological or genetic analysis methods. We do observe evidence of coincidence at some individual genetic loci (such as *PPP1R15A, TYK2*) between SARS-CoV-2 infection and COVID-19 severity related variants and blood cell related variants. We are hopeful that as more balanced case-control studies with increased sample sizes become available for COVID-19 related phenotypes more definitive conclusions regarding the similarity of blood cell count and COVID-19 genetic architecture can be raised.

## Methods

We first examined the relationship between measured blood cell indices and COVID-19 related outcomes in two biobank cohorts. Next, we performed analyses assessing coincidence between GWAS identified loci associated with SARS-CoV-2 infection or COVID-19 severity and loci identified for blood cell traits. Finally, in order to examine the shared genetic architecture of COVID-19 and hematological traits, we performed a genetic correlation analysis with LD Score Regression [30] and a MR analysis with the Inverse Variant Weighted estimator [58], with MR-Egger [59] and the weighted median estimator also performed as sensitivity analyses. We analyzed publicly available summary statistics for three COVID-19 phenotypes from the HGI: COVID-19 severity as measured by severe respiratory infection (phenotype A2), COVID-19 severity as measured by hospitalization (phenotype B2), and SARS-CoV-2 infection (phenotype C2) [29]. For hematological traits, we utilized summary statistics from the largest GWAS to date of 408,112 European ancestry participants in the UKB cohort [23]. We note that all genomic positions throughout this manuscript are from build 37, and all LD calculations, in text and figures, are from TOP-LD [60] unless otherwise noted.

### Data

#### UK Biobank (UKB) Samples from Measured Blood Trait analyses

UKB (http://www.ukbiobank.ac.uk/resources/) recruited 500,000 people aged between 40-69 years in 2006-2010, establishing a prospective biobank study to understand risk factors for common diseases such as cancer, heart disease, stroke, diabetes, and dementia). Participants are being followed-up through health records from the UK National Health Service. UKB has genotype data on all enrolled participants, as well as extensive baseline questionnaires and physical measures and stored blood and urine samples. Hematological traits were assayed as previously described [21]. Genotyping on custom Axiom arrays and subsequent quality control has been previously described [61]. Samples were also excluded based on factors likely to cause major perturbations in hematological indices. For example, we dropped samples based on positive pregnancy status, certain drug treatments, cancer self-report, ICD9 and ICD10 disease codes, and surgical procedures (**Supplementary Table 6**), as well as individuals who have withdrawn consent. Samples were included only if they had complete data for all covariates and phenotypes (n= 423,358).

##### UKB Summary Statistics

Summary statistics were obtained from a recent GWAS of hematological traits in UKB participants of European ancestry [23]. As a brief overview of the analysis, a GWAS was conducted on 29 hematological traits in European ancestry participants in the UKB. Raw phenotypes were regressed on age, age-squared, sex, principal components and cohort specific covariates (e.g., study center, cohort, *etc*), and WBC-related traits were log10 transformed before regression modeling. Residuals from the modeling were obtained and then inverse normalized for cohort level association analysis or GWAS.

#### Vanderbilt University Medical Center, clinical cohort samples

Vanderbilt University Medical Center is a major comprehensive and tertiary care center in Nashville, Tennessee. The Synthetic Derivative (SD), a completely de-identified copy of the electronic health record (EHR), contains longitudinal clinical information for over 3.3 million individuals [62]. The database incorporates information from multiple sources and includes diagnostic and procedure codes (International Classifiers of Disesase [ICD] and Current Procedural Terminology [CPT], respectively), demographics (age, gender, EHR-reported race and ethnicity), text from clinical care (i.e. discharge summaries, nursing notes, progress notes, history and physical, problem lists), medications, and laboratory values. The QualityLab pipeline was used to extract and clean laboratory values for over 275 million observations across 1.5 million patients as previously described [63].

#### HGI COVID-19

We downloaded the round 5 (January 18, 2021) COVID-19 summary statistics for three phenotypes for European ancestry participants from https://www.covid19hg.org. The critically ill phenotype was defined included patients who were hospitalized due to symptoms associated with laboratory-confirmed SARS-CoV-2 infection and who required respiratory support or whose cause of death was associated with COVID-19 (5,101 cases and 138,3241 controls). The hospitalization phenotype is a binary indicator of patients who were hospitalized for symptoms associated with laboratory-confirmed SARS-CoV-2 infection (9,986 cases and 1,877,672 controls). Population controls were used in both cases, including individuals whose exposure status to SARS-CoV-2 was unknown. Both phenotypes were defined using diagnostic criteria following the Diagnosis and Treatment Protocol for Novel Coronavirus Protocol [64]. Individuals with positive reported infection status were compared against population controls (38,984 cases and 1,644,784 controls).

We note that there is a more recent COVID-19 meta-analysis release (Round 6, June 15, 2021) but this release did not include versions of the summary statistics limited to European populations or excluding UKB, which were necessary for some of the analyses contained here to avoid bias due to sample overlap (particularly for MR-Egger[59]).

### Measured Blood Cell Analysis

Blood cell indices were directly measured using blood draws from the UKB in-person visits, as described above. We assessed the association of each measured blood cell trait with SARS-CoV-2 infection status (positive test versus population) and with COVID-19 hospitalization (any hospitalized case versus population). Logistic regression analyses for case/control analyses were adjusted for age, sex, assessment center, self-reported ethnicity (indicator variables for “White”, “Mixed”, “Asian or Asian British”, “Black or Black British”, or “Chinese”, based on Data-Field 21000) time elapsed between blood cell trait measurement and either SARS-CoV-2 test date or date of data download, and a blood cell trait and time elapsed interaction term. All data was downloaded on March 12, 2021, for the analyses presented here. SARS-CoV-2 test results were available through February 24, 2021, including location of test, and hospitalization related data were available through March 7, 2021. Cause of death data was available through February 16, 2021. For consistency with data available in VUMC SD, we included as a hospitalized case anyone with a positive SARS-CoV-2 test originating when they were an inpatient at a hospital, or with a positive test up to 14 days before hospital admission, during hospital episode, or within 7 days after hospital discharge. This definition would not exclude cases identified incidentally in individuals hospitalized for other reasons. Controls are defined as the rest of the population, for consistency with other genetic analyses [24].

### VUMC SD Replication

We also assessed association of measured blood cell traits with SARS-CoV-2 positive test status and COVID-19 hospitalization in the VUMC SD [65]. Lab values were extracted from de-identified medical records and cleaned as previously described, using the QualityLab pipeline [63]. At least 50 cases were required for a model to be tested. Due to low case counts for COVID-19 outcomes, analyses were performed in a multiracial background with EHR-reported race and ethnicity used as covariates in the models. Case and control populations were extracted from the SD by experienced programmers with data current to October 2020. A SARS-CoV-2 positive test was determined by either a positive SARS-CoV-2 test or related diagnostic code, U07.1. Controls included all individuals with data for a given blood cell trait and absence of any SARS-CoV-2 positive test. Cases for the COVID-19 hospitalization analysis are individuals with a SARS-CoV-2 positive test and a hospitalization code (for any indication) in the seven days before or 30 days after a positive SARS-CoV-2 test. Controls for the COVID-19 hospitalization analysis are similarly defined as all individuals with blood cell trait data that do not meet the case definition; however, we note that we also performed a sensitivity analysis using as controls individuals with a positive SARS-CoV-2 test and no hospitalization code within a similar time window (with similar results, not shown).

Similar to UKB analysis, replication analyses included logistic regressions adjusted for age, sex, EHR-reported race and ethnicity, time elapsed between blood cell trait measurement and either SARS-CoV-2 test date or end of follow-up, and a blood cell trait and time elapsed interaction term. As is commonly done in biobank data, median blood cell trait values were used across all available timepoints, with time of follow-up calculated in reference to the median age at which a given blood cell measurement was obtained. VUMC SD and UK Biobank results were combined using fixed effects meta-analysis.

### Analysis of Coincident Genetic Association Signals

We assessed evidence for coincident genetic association signals at COVID-19 loci which had been reported as associated with blood cell traits in a previous GWAS [23, 29]. We used the European TOP-LD [60]reference panel to establish COVID-19 loci in LD (r^2^ > 0.4) with distinct blood cell trait variants. To ensure that our results were not contaminated by sample overlap, at each COVID-19 locus from the full HGI meta-analysis results, we used the lead variant in the results excluding UKB as a proxy for analysis of coincident signals.

### Genetic Correlation

LD Score Regression is a statistical technique to estimate genetic correlation between complex traits using only GWAS summary statistics [30]. We obtained LD scores from the publicly available HapMap3 weights, and restricted our analysis to only consider 1,217,090 HapMap3 SNPs [30]. We estimated genetic correlation for each combination of blood cell trait and COVID-19 phenotype. Statistical significance was evaluated as nominally significant at a p-value less than 0.05, and Bonferroni significant using 0.05/87. Pairs of traits which demonstrated at least nominal statistical significance of non-zero genetic correlation were prioritized in the MR analysis.

### Mendelian Randomization (MR)

MR was performed in order to estimate the causal effect of hematological traits on the three COVID-19 phenotypes. 16,900 conditionally independent associations previously identified in UKB participants of European ancestry were used as instruments for the hematological traits[23]. Briefly, these distinct variants were determined as conditionally independent signals via stepwise multiple regression. The IVW estimator served as our primary estimator of the causal effect. As a sensitivity analysis, we fit the MR-Egger model in order to assess the evidence of directional pleiotropy via the MR-Egger intercept test. If the MR-Egger test demonstrated moderate statistical evidence away from the null hypothesis that it is zero (p < 0.2), we report the MR-Egger estimate of the causal effect rather than the IVW estimator. We further estimated the weighted median causal effect for robustness.

## Data Availability

For the measured blood cell analyses, due to participant privacy individual level data cannot be shared but is available upon request from UK Biobank. Genetic summary statistics are available for download at ftp://ftp.sanger.ac.uk/pub/project/humgen/summary_statistics/UKBB_blood_cell_traits/ (for blood cell traits) and https://www.covid19hg.org/results/r5/ (for COVID-19 related phenotypes).

ftp://ftp.sanger.ac.uk/pub/project/humgen/summary_statistics/UKBB_blood_cell_traits/

https://www.covid19hg.org/results/r5/

## Abbreviations

GWAS: Genome-wide association study
HGI: COVID-19 Host Genetics Initiative
UKB: UK Biobank
LD: linkage disequilibrium
RBC: red blood cell
RDW: red blood cell distribution width
MCH: mean corpuscular hemoglobin
IRF: immature reticulocyte fraction
1000G: 1000 Genomes Project Phase 3 v5
MR: Mendelian randomization
IVW: inverse-variance weighted
VUMC: Vanderbilt University Medical Center
SD: Synthetic Derivative

## Declarations

### Ethics Approval

UK Biobank has approval from the North West Multi-centre Research Ethics Committee (MREC) as a Research Tissue Bank (RTB) approval. VUMC SD has been approved by the Vanderbilt University IRB and the Operations Oversight Board (OOB).

### Conflict of Interest Statement

The authors declare that they have no competing interests.

### Funding

This research was funded by R01HL146500 and U01HG011720. The project described was also supported by the National Center for Advancing Translational Sciences, National Institutes of Health, through Grant KL2TR002490 (LMR).

The SD projects at Vanderbilt University Medical Center are supported by numerous sources: institutional funding, private agencies, and federal grants. These include the NIH funded Shared Instrumentation Grant S10OD017985 and S10RR025141; CTSA grants UL1TR002243, UL1TR000445, and UL1RR024975 from the National Center for Advancing Translational Sciences. Its contents are solely the responsibility of the authors and do not necessarily represent official views of the National Center for Advancing Translational Sciences or the National Institutes of Health. Genomic data are also supported by investigator-led projects that include U01HG004798, R01NS032830, RC2GM092618, P50GM115305, U01HG006378, U19HL065962, R01HD074711; and additional funding sources listed at https://victr.vumc.org/biovu-funding/.

## Acknowledgements

Support for title page creation and format was provided by AuthorArranger, a tool developed at the National Cancer Institute.

This research has been conducted using the UK Biobank Resource under Application Number 25953. We would like to thank the Covid-19 Host Genetics Initiative for sharing the results of their analyses.

## Authors’ contributions

BR performed the analysis and drafted the manuscript. QS, WW, TMF, MMS, AF, MG, and LMR also performed analyses or derived critical phenotypes. NC, EEB, and PA helped conceptualize the project and contribute to manuscript drafting. YL, VGS, APR and LMR supervised the work, conceptualized the project, and contributed to manuscript drafting.

All authors read and approved the final manuscript.

## Supplementary Figure

**Figure S1.**
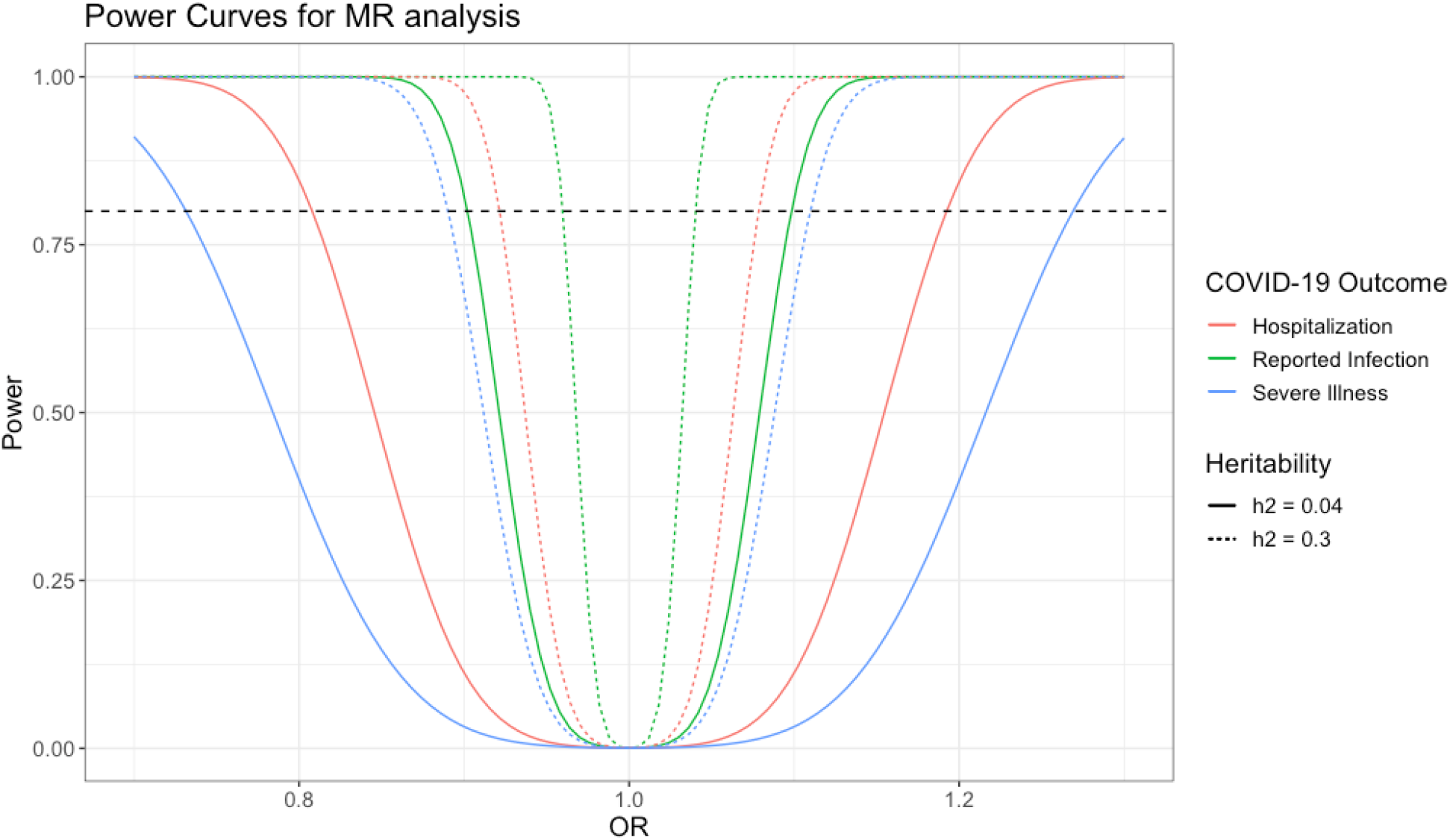
Power curves for Mendelian Randomization analysis, based on round 5 HGI meta-analysis sample sizes [24]. Legend: h2 = h^2^.

## Notes

### Competing Interest Statement

The authors have declared no competing interest.

